# Prevalence of Hypertension in a STEMI-activated Subset of Patients from 2019 to 2021

**DOI:** 10.1101/2023.09.20.23295873

**Authors:** Trang Nguyen, Deborah Lee, Rebekah Lantz, Dylan Hefner, Casey T Walk

**Affiliations:** Molecular Medicine., 3640 Colonel Glenn Hwy, Fairborn, OH 45324; Biology, Korean., 3640 Colonel Glenn Hwy, Fairborn, OH 45324; 1 Wyoming St, Dayton OH 45409; Applied Health Science., 3640 Colonel Glenn Hwy, Fairborn, OH 45324; Pharmacology and Toxicology., 3640 Colonel Glenn Hwy, Fairborn, OH 45324

**Keywords:** STEMI, STEMI-activated, COVID-19, Hypertension, Cardiovascular disease, Coronary Risk Factor, Pandemic, Stroke belt, Regional, Population health, Compliance, Antihypertensive, Ohio

## Abstract

**Background:** About 47% of United States adults have hypertension (HTN) per the 2017 Hypertension Clinical Practice Guidelines, where the blood pressure target is <130/90 mmHg. Prevalence is worse in certain distributions of the country with top quartile in the Southeastern pocket. In a subset of patients who present with ST-segment elevated myocardial infarction (STEMI), the prevalence is 30-40%. An interesting further subset of patients are those who presented as STEMI-alert during the COVID-19 pandemic. COVID-19 wreaked havoc on the world with its first confirmed case in 9 January 2020. International followed by national transport became limited, followed by announcement of a pandemic. Quarantines and lockdowns were set in place, especially in March 2020, to prevent epidemiologic spread.

**Hypothesis:** We were interested to observe if STEMI-alert patients during this period presented with worse background disease compared to years 2019 and 2021, years before and after COVID-lockdown and peak, including if the prevalence of background HTN increased in the population.

**Methods:** We evaluated 1001 adults who were STEMI-activated from 1 January 2019 to 31 December 2021 at five sites in Southwest Ohio. We obtained patient demographics and risk factors and performed multinomial logistic regression to compare years 2019 and 2021 to 2020. Statistical analysis was with SAS 9.4.

**Results:** For 1001 STEMI-alert patients, 244 patients (72.6%) had HTN in 2019, 250 (78.9%) in 2020 and 261 (75.0%) in 2021. Overall prevalence over the three years was 755 (75.4%). Compared to 2020, 2019 prevalence was not significant (p=0.12) (OR 0.72) (CI 0.47,1.09). Neither was 2021 (p=0.25) (OR 0.78) (CI 0.51,1.19).

**Conclusion:** STEMI-alert patients at our institution appeared to have higher overall prevalence of HTN than reported nationally by the Centers for Disease Control (CDC), 75.4% versus 47%. These may have self-selected for disease severity by STEMI-alert activation status. HTN may also be of higher prevalence in this region, be associated with poor disease detection or there may be patient compliance issues when antihypertensives are prescribed by a healthcare provider. HTN prevalence did not have statistical significance across the three observed years despite our hypothesis that patients would present with more background cardiac comorbidities such as HTN.

## Introduction

With the first confirmed case of COVID-19 on 9 January 2020, the world had changed. Global air travel was banned in February and the World Health Organization (WHO) declared COVID-19 a pandemic in March 2020. National travel also ceased, along with quarantine and lockdowns enacted to prevent epidemiological spread.^1^ Twelve percent of all deaths between March of 2020 and October of 2021 in the United States (US) were attributed to COVID-19, making it the third leading cause of death after heart disease and cancer.^2^ COVID-19’s impact was significant to cause categorization of the world timeline from before COVID-19 to afterwards for many comorbidities and primary diseases.

From 2019 to 2020, deaths due to heart disease rose by 4.1%.^3^ There was a 10-times increase in mortality for individuals with pre-existing cardiovascular comorbidities than those without, along with higher secondary heart injury treatment complications in patients positive for COVID-19.^4,5^ Comorbidities noted to increase risk continue to include diabetes, hypertension (HTN), and chronic lung disease.^6^ In patients with COVID-19, there was a higher prevalence of hypertension, diabetes mellitus, and cardiovascular disease. Additionally, direct cardiac manifestations of COVID-19 can be a contributing factor of patient mortality. These cardiovascular manifestations include myocardial injury (MI) due to inflammation, nonischemic cardiomyopathy, pericarditis, coronary vasospasm, and stress cardiomyopathy.^6^

Early outcome data from the US and China showed HTN to be among the most prevalent cardiovascular related comorbidities in COVID-19 patients. Hypertensive patients were more likely to be admitted to the hospital.^7^ According to the 2017 Hypertension Clinical Practice Guidelines where HTN is diagnosed for blood pressure ≥130/90 mmHg, 47% equivalent to 122.4 million, of US adults ≥20 years old have HTN.^8^ HTN is strongly associated with cardiovascular and chronic kidney disease risk and increased overall mortality.^9^ The burden of HTN has been increasing with an estimated 10% of global healthcare expenses attributed to its care.^9^

In patients with ST-segment elevated myocardial infarction (STEMI), the prevalence of background HTN is 30-40%.^10^ In addition, a population of interest is the era of COVID-19 in relation to HTN and MIs. Amid the COVID-19 pandemic, many hospitals canceled all in-person elective patient care, shifting to telemedicine options where able. Among practices that were canceled, routine checkups were included, conceivably impacting the management of chronic illnesses such as HTN.^7^ With information we know about COVID-19 and HTN, we were interested to observe if STEMI-alert patients during this lockdown and peak cases period in 2020 presented with worse background disease compared to 2019 and 2021. We were also interested to see if the prevalence of background HTN increased with time trends overall in our three-year retrospective study. The abstract for this study was previously presented at American Heart Association Hypertension 2023.^11^

## Methods

This observational study was exempted by Wright State University Institutional Review Board (#07224) and included 1001 patients who were STEMI-activated between 1 January 2019 through 31 December 2021, to include a sample of patients pre, peri, and post COVID-19 peak cases and lockdown. Patients were included if they presented to any of five inpatient sites within a hospital system in Southwest Ohio, were aged 18 years or older, and STEMI-activated upon presentation, either en route to the hospital, interfacility transfer, or in-hospital stay. Patients were excluded if they were <18 years old, were considered vulnerable persons per the Declaration of Helsinki Ethical Principles,^12^ or were not a STEMI-activation. Duplicate entries were removed and favored the index event.

Seven categories of interest were gathered and evaluated from Epic electronic medical record (EMR) using Microsoft SQL Server Manager and from National Cardiovascular Data Registry (NCDR) to determine if there were significant differences in various patient characteristics between 2019 through 2021. They included baseline characteristics, details of presentation, procedure details, diagnostic values, rationale, time variables, and outcomes. Data were analyzed with SAS version 9.4 (SAS Institute, Inc., Cary, NC) and RStudio version 2022.07.1 (RStudio, Inc., Boston, MA). A level of significance of alpha = 0.05 was used throughout to assess statistical significance.

A multinomial logistic regression analysis was conducted to examine the relationship between baseline characteristics and the years 2019, 2020, and 2021. The response variable was defined as the year, while the predictor variables included relevant parameters such age, gender, body mass index (BMI), hypertension, dyslipidemia, diabetes mellitus, cerebrovascular disease, peripheral arterial disease, chronic lung disease, prior coronary artery bypass graft, tobacco use, current dialysis status, congestive heart failure, and newly diagnosed heart failure. The year 2020 was designated as the reference level for comparison given its central importance. Consequently, the focus of the analysis was on discerning significant disparities or correlations between the variables of interest in 2020 relative to 2019 and 2021. This model enables the estimation of the odds pertaining to a patient’s likelihood of presenting with a specific baseline characteristic in the years 2019 or 2021 in comparison to the reference year.

## Results

The study encompassed a cohort of 1001 patients who underwent STEMI activation, with 336 cases in 2019, 317 in 2020, and 348 in 2021. The mean age at presentation was 63.5 years, the average body mass index (BMI) was 30.2 kg/m^2^, and the mean preprocedural ejection fraction was 51.3%. The independent predictor for outcomes was the year of presentation. The full details of patient presentation and outcomes are previously published in Journal of Clinical Cardiology and Cardiovascular Interventions.^13^ Outcomes are separately presented and published in CHEST.^14^

Figure 1 shows results of HTN in this STEMI-activated cohort during the observed COVID-19 era. Across the study duration, a total of 755 (75.4%) patients who presented with STEMI-activation had background hypertension. This was partitioned by year and it was found that 244 (72.6%) patients in 2019, 250 (78.9%) in 2020, and 261 (75.0%) in 2021 had background HTN [Figure 1]. Compared to 2020, 2019 did not have statistical significance for hypertensive patients (p=0.12). The odds ratio (OR) for this comparison was 0.72, accompanied by a confidence interval (CI) of 0.47 to 1.09. Similarly, when comparing the prevalence between 2020 and 2021, the observed difference lacked statistical significance (p=0.25), with an OR 0.78 and a CI of 0.51 to 1.19. Thus, the observed trends in the prevalence of hypertension over the three-year span did not exhibit statistically significant variations between respective years [Table 1].

**Table 1.** Comparative values for hypertensive cases compared to 2020.

| Table 1. Comparative values for hypertensive cases compared to 2020. |  |  |  |  |
| --- | --- | --- | --- | --- |
| Year | Hypertensive cases, N (%) | p-value | OR | CI |
| Overall | 755 (75.4%) |  |  |  |
| 2019 | 244 (72.6%) | p=0.12 | 0.72 | 0.47,1.09 |
| 2020 | 250 (78.9%) |  |  |  |
| 2021 | 261 (75.0%) | p=0.25 | 0.78 | 0.51,1.19 |
| Definitions: CI, confidence interval. OR, odds ratio. |  |  |  |  |

## Discussion

The findings of our study elucidate the prevalence of HTN among STEMI patients over a three-year period, from 2019 to 2021. Hypertension continues to be prevalent and has strong associations with cardiovascular disease and overall mortality.^9^ Thus, understanding its prevalence in this specific patient population is of great clinical significance.

Our analysis is consistent with previous literature by illustrating that hypertension is indeed an impactful comorbidity for cardiovascular disease. Fuchs and Whelton described that high blood pressure has the strongest association with the causation of cardiovascular disease.^15^ These studies align with our results that HTN plays a large role in cardiovascular disease and STEMI-MIs.

A systematic review of PubMed, Web of Science, and EBSCOHost (MEDLINE) that included 7545 hypertensive emergency patients to study the prevalence of MI in hypertensive crises was undertaken by Talle et al. to learn more about HTN and MI associations. Fifteen (83.3%) of the studies indicated MI prevalence with a wide range from 3.6-59.6% and 3 studies (16.7%) indicated MI prevalence of 15-63%, which still remains a wide range. Overall, not many studies in prior literature have focused on MI among patients with background HTN, hypertensive emergencies, and its relation to MI, indicating an increased need for research to provide important data and assist with management of HTN and to decrease risk for adverse cardiovascular effects.^16^

We also considered the impact of COVID-19 on HTN control, where patients were encouraged to stay home to reduce risk of viral spread during a lockdown period in 2020. Shah et al. observed blood pressure home readings before and during COVID-19.^17^ The analysis enrolled 72,706 patients via a digital hypertension program between January 2019 to March 2020 and April 2020 to August 2020. Results showed the mean monthly home blood pressure, including the adjusted mean systolic blood pressure, diastolic blood pressure and mean arterial pressure, increased during COVID-19, 2020, compared to pre-COVID, 2019.^17^ The increase in blood pressure could be due to the change in clinical practices during the pandemic.

A result that differed from the literature was STEMI-alert patients at our institution appeared to have higher overall prevalence of background HTN than reported nationally, 75.4% in our data compared to 47% reported nationally by the Centers for Disease Control (CDC) according to the 2017 Hypertension Clinical Practice Guidelines.^8^ A possible rationale for this discrepancy is that the study may have self-selected for disease severity by STEMI-alert activation status. However, our figure remains higher than 30-40% from the STEMI-subset of patients noted in The Journal of Clinical Hypertension published in 2019.^10^ Patients who experience more severe symptoms or are at higher risk for cardiac events may be more likely to activate the STEMI-alert at an institutional level, leading to a higher prevalence of HTN within this subgroup. We suspected a regional component in our Southwest Ohio facilities. Regional variations in disease prevalence are well-documented from the Coronary Artery Risk Development in Young Adults (CARDIA) study and higher prevalence is noted in the Southeastern region of the US, also termed the Stroke Belt.^18^ Similarly modifiable and nonmodifiable risk factors that lead to stroke also lead to cardiac events including lifestyle, genetics, and healthcare access.^18^ Further investigation into regional patterns of HTN may provide valuable insights, which is one of the benefits of our study and can benefit from replication.

The high prevalence of HTN among STEMI-alert patients at our institution remained consistent throughout the study period. In 2019, 72.6% of STEMI patients were diagnosed with background HTN, which increased to 78.9% in 2020, and slightly decreased to 75.0% in 2021 without a statistical significance in these differences. The overall prevalence over these three years as 75.4% before and after the COVID-19 peak cases was unexpected. Literature supports that the most prevalent comorbidities of COVID-19 patients were underlying HTN, cardiovascular disease, and diabetes, with a higher HTN prevalence in patients with more adverse outcomes.^19^ Thus, we hypothesized that there may be worse background disease in STEMI patients during the peak of COVID, but the results were contrary and may benefit from a longer trend over time.

The results of no significant HTN trend between the years could be due to the baseline high regional HTN, as mentioned above. This absence of significance could also be due to other complex factors that play a role in STEMI patients during the COVID-19 era that makes isolating the role of just one factor, HTN, difficult. Literature indicates that age, sex, family history, cholesterol levels, tobacco use, obesity, and diabetes are a few risk factors for STEMI. Concordantly, advanced age, positive smoking status, obesity, elevated D-dimer, and chronic kidney injury are a few risk factors for worser manifestation of COVID-19 and adverse outcomes including death.^20-21^ Thus the role of HTN in STEMI and cardiovascular outcomes during COVID may be mixed with other risk factors. It is also worth considering the possibility of patient compliance issues with antihypertensive medications prescribed by healthcare providers.

Patients had limited access to healthcare during the pandemic, which may have included refilling prescription medications.^7^ Non-adherence and limited access to treatment can contribute to uncontrolled HTN, and addressing this issue may be essential in reducing the burden of HTN-related complications in STEMI patients and generally.

Strengths of our study include that to the best of our knowledge, the topic of HTN in a subset of STEMI-activated patients during COVID-19 has not been thoroughly studied and thus the topic of our study is unique. In addition, we may have found an area for quality improvement in the Southwest Ohio area as the results indicate higher-than-US-average prevalence of HTN. Regarding strength of our study design, the longitudinal design spanning three consecutive years allows for analyzing trends in HTN prevalence among STEMI patients over time. The study’s inclusion of 1001 patients increased the power of the analysis. Given that it was conducted retrospectively in a real-world clinical setting, the results are likely to reflect the genuine prevalence of HTN in this specific patient population, offering direct clinical relevance.

Some of the limitations of our research pertains to its geographic scope, exclusively focused on a Southwest Ohio region. Data was collected from five hospitals in a single healthcare system, potentially limiting the generalizability of the findings which may not account for regional variations observed elsewhere. Future investigations could explore different geographical regions to broaden the study’s applicability. The reliance on healthcare system data also means that certain patient characteristics and lifestyle factors may not be fully captured by the billed diagnoses and documented history that were collected during index encounters. The study recognizes the possibility of selection bias, as STEMI-activated patients may differ from their non-STEMI-activated counterparts in terms of disease severity and risk factors, which could influence the observed prevalence of HTN. Additionally, HTN and its negative outcomes typically require an extended period to manifest, making it challenging to accurately interpret data within shorter timeframes. Despite the study’s three-year duration, the lack of statistical significance for changes in HTN prevalence over time raises questions about its ability to detect meaningful differences and calls for further investigation into potential contributing factors.

In conclusion, we found that 75.4% of our STEMI cohort presented with HTN, averaging from 72.6% in 2019, 78.9% in 2020, and 75.0% in 2021. Our analysis indicated that there was no statistically significant variation between the prevalence of HTN between the three years. Our study results are consistent with literature emphasizing HTN as an important comorbidity with cardiovascular disease and COVID-19. However, our cohort’s background HTN prevalence was much higher than the national mean, 75.4% versus 47%, and 75.4% versus 30-40% for those STEMI-activated presentations. These findings underscore the importance of tailored management and interventions for this subgroup of patients. Further research is needed to explore the underlying reasons for this high prevalence, including regional factors and patient behaviors, and to develop strategies aimed at improving HTN control in this population. Understanding these dynamics is critical in enhancing the overall care and reducing adverse outcomes of STEMI patients with HTN.

## Data Availability

Please contact the corresponding author.

